# Effects of cognitive behavioral therapy for insomnia on subjective and objective measures of sleep and cognition

**DOI:** 10.1101/2021.10.21.21265019

**Authors:** Aurore A. Perrault, Florence B. Pomares, Dylan Smith, Nathan E. Cross, Kirsten Gong, Antonia Maltezos, Margaret McCarthy, Emma Madigan, Lukia Tarelli, Jennifer McGrath, Josée Savard, Sophie Schwartz, Jean-Philippe Gouin, Thien Thanh Dang-Vu

**Affiliations:** Sleep, Cognition and Neuroimaging Lab, Department of Health, Kinesiology and Applied Physiology & Center for Studies in Behavioral Neurobiology, Concordia University, Montreal, Quebec, Canada; Centre de Recherche de l’Institut Universitaire de Gériatrie de Montréal, CIUSSS Centre-Sud-de-l’Ile-de-Montréal, Québec, Canada; Stress, Interpersonal Relationship and Health Lab, Department of Psychology & Centre for Clinical Research in Health, Concordia University, Montreal, Quebec, Canada; Sleep Unit, University of Ottawa Institute for Mental Health Research, Ottawa, Ontario, Canada; Pediatric Public Health Psychology Laboratory, Department of Psychology & Centre for Clinical Research in Health, Concordia University, Montreal, Quebec, Canada; School of Psychology, Université Laval and CHU de Québec-Université Laval Research Center, Québec, Canada; Department of Neuroscience, Faculty of Medicine and Swiss Center for Affective Sciences, University of Geneva, Geneva, Switzerland; PERFORM center, Concordia University, Montreal, Quebec, Canada

**Keywords:** CBTi, sleep, cognition, objective, subjective, sleep-state misperception

## Abstract

**Study Objectives:** To assess the effects of Cognitive Behavioral Therapy for insomnia (CBTi) on subjective and objective sleep, sleep-state misperception as well as self-reported and objective cognitive performance.

**Methods:** We performed a randomized controlled trial with a treatment group and a wait-list control group to assess changes in insomnia symptoms after CBTi (8 sessions/3 months) in 62 participants with chronic insomnia. To this end, we conducted a multimodal investigation of sleep and cognition including subjective measures of sleep difficulties (Insomnia Severity Index (ISI), sleep diaries) and cognitive functioning (Sahlgrenska Academy Self-reported Cognitive Impairment Questionnaire), objective assessments of sleep (polysomnography recording, cognition (attention and working memory tasks), and sleep-state misperception measures, collected at baseline and at 3-months post-randomization. At 6 months post-randomization, we collected similar data from the wait-list group after CBTi. We also assessed ISI one year after CBTi in both groups. Our main analysis investigated changes in sleep and cognition after 3 months (treatment versus wait-list group). In secondary analyses, we pooled data from both groups to observe changes after CBTi.

**Results:** ISI score was reduced and self-reported sleep quality improved after CBTi (treatment group at 3 months and pooled groups after CBTi). Sleep misperception in sleep onset latency and sleep duration decreased after CBTi. In contrast, objective sleep, objective and subjective cognitive functioning did not improve after CBTi.

**Conclusions:** We showed that CBTi has a beneficial effect on variables pertaining to the subjective perception of sleep, which is a central feature of insomnia. However, we observed no significant effect of CBTi on measures of cognitive functioning.

**STATEMENT OF SIGNIFICANCE:** Nighttime sleep difficulties and daytime cognitive impairment are the two main complaints of individuals suffering from insomnia. We investigated the effects of Cognitive Behavioral Therapy for insomnia (CBTi) on these two aspects of chronic insomnia, using both self-reported and objective measures, as well as sleep-state misperception (i.e., the discrepancy between self-reported and objective sleep). We showed significant changes after CBTi in insomnia severity, subjective sleep quality and perception of sleep, while no consistent benefit emerged for objective measures of sleep and assessments of cognitive functioning. CBTi thus appears to primarily benefit subjective sleep quality as well as the alignment between subjective and objective estimates of sleep.

## INTRODUCTION

Chronic insomnia is defined by self-reported complaints of difficulty falling asleep and/or maintaining sleep, occurring at least 3 times per week for more than 3 months, and is accompanied by daytime functioning complaints^1^, such as fatigue, lack of energy, mood disruption, and often, cognitive complaints^2,3^. Chronic insomnia affects more than 10% of the population^4–6^. It is associated with poorer physical and mental health outcomes and an important societal financial burden^7–9^, thereby representing a major health issue. Current pharmacotherapeutic treatments are associated with greater risk of tolerance, dependence, drug abuse^10,11^. Given its long-term efficacy, the first-line of treatment for chronic insomnia is cognitive-behavioral therapy for insomnia (CBTi)^12,13^, a multimodal psychological intervention aimed at modifying maladaptive thinking and behaviors that contribute to the perpetuation of insomnia^14,15^.

The effectiveness of CBTi on insomnia severity has been extensively investigated. Previous studies have shown CBTi to be effective on insomnia severity and subjective sleep quality, with medium to large effect sizes^12,16–19^ using mostly self-report questionnaires, such as the Insomnia Severity Index (ISI)^20^ or the Pittsburgh Sleep Quality Index (PSQI)^21^. Sleep diaries, collected daily over several days provide a comprehensive assessment of self-reported insomnia symptoms^22^. CBTi has been shown to result in diary-reported improvements in sleep initiation (i.e., shortened sleep onset latency) and maintenance (i.e., shorter wake after sleep onset) as well as sleep satisfaction (i.e., sleep quality, sleep efficiency) with moderate to large effect sizes^12,23,24^. These beneficial effects on self-reported outcomes have been observed immediately after the completion of CBTi^12^ and at longer term (between 6 months and several years post-CBTi)^23,24^. However, in these studies, the changes on self-reported sleep duration remained small to non-significant.

While the efficacy of CBTi has been consistently confirmed with self-reported (i.e., subjective) outcomes, concomitant changes in objective sleep measures have not been systematically investigated. Polysomnographic (PSG) sleep studies revealed conflicting results on the effects of CBTi on objective sleep outcomes, exhibiting either small effect sizes or no improvement^12,24^. As insomnia is a subjective disorder for which the diagnosis is solely based on subjective complaints, such observations suggest that CBTi is effective in improving subjective sleep quality rather than actively changing sleep architecture per se. Notably, the subjective sleep complaints reported by individuals with chronic insomnia are not always corroborated by objective sleep measures, such as polysomnographic (PSG) assessment^25^. Such discrepancy between self-reported and objective sleep assessments can be quantified by measures of sleep-state misperception, i.e., differences between subjective and objectives estimates of sleep^26^. Some individuals with chronic insomnia underestimate their sleep duration and overestimate their sleep latency when comparing their reports to objective sleep measures^27,28^. Few studies reported the effects of insomnia treatment on sleep-state misperception (SSM) with actimeter^29,30^ and only one study compared PSG to self-reported sleep measures after CBTi^31^. Lund and collaborators (2013) found less misperception of sleep onset latency (i.e., reduced discrepancy) without change in objective sleep in a sample of older adults with insomnia comorbid with osteoarthritis, coronary artery disease or chronic obstructive pulmonary disease. These results suggest that, in addition to self-reported sleep quality, CBTi also improves the perception of sleep. These findings await replication in broader samples of individuals with chronic insomnia.

Beyond nighttime effects, CBTi has been shown to also improve quality of life and daytime functioning^3,32^. The effectiveness of CBTi on daytime functioning – including greater energy, better concentration and mood – has shown a large effect size compared to a wait-list control, and a small effect size compared to a placebo intervention^32^. With regards to cognitive function, the effect of CBTi on subjective cognitive function has been reported small to moderate and not specific to one cognitive domain^33^. Findings are inconsistent for objective cognitive measures; some studies reported an improvement in attentional abilities after CBTi, with improved objective measures of attention in the Attentional Network Test-Interactions^34^, while other studies reported null findings in neuropsychological and vigilance tasks^35,36^. While no study has reported the effects of CBTi on both subjective and objective cognitive performances using standardized in-person intervention and assessments, one study investigated the effect of CBTi on both subjective and objective cognitive abilities, using web-based intervention (digital CBTi) and assessments^19^. Improvement following digital CBTi were only found on self-reported cognitive functioning using the British-Columbia Cognitive Complaints Inventory questionnaire. However, objective cognitive performance across multiple cognitive domains (attention, memory, mental flexibility, complex processing speed) assessed using the cognitive test battery from the UK Biobank did not improve^19^, confirming previous results^33^ but also the crucial subjective dimension of chronic insomnia.

In sum, while the effects of CBTi on subjective and objective sleep are well documented separately, there is limited data about its effects on sleep-state misperception (i.e., discrepancy between objective and subjective sleep) and on subjective and objective measures of cognitive functioning. Here, we aimed at providing a more comprehensive assessment of the effects of CBTi on sleep and cognitive function within the same group of individuals. Specifically, we conducted a multimodal investigation of CBTi efficacy on sleep and cognition using combined objective and subjective assessments in a randomized-controlled trial with 2 arms (i.e., immediate CBTi and wait-list control) on 62 participants with chronic insomnia. The ISI was used as our primary outcome of CBTi efficacy. Secondary outcomes included subjective (self-report) measures of sleep difficulties (i.e., sleep diaries) and subjective cognitive functioning (i.e., questionnaires), as well as objective assessments of sleep (i.e., PSG recording), cognition (i.e., attention and working memory tasks) and sleep-state misperception measures of sleep duration and sleep latency. Both primary and secondary outcomes were assessed at baseline and after 3 months (after immediate CBTi or after wait-list period). The wait-list control group then received the CBTi program and outcomes were again assessed after CBTi (i.e., 6 months post-randomization). We pooled both groups Post-CBTi for complementary analyses of CBTi efficacy on sleep and cognition. We hypothesized that CBTi would have a large effect on subjective (self-reported) measures of sleep, a small-to-medium effect on subjective cognitive functioning, while objective measures of sleep and cognition would yield small to no changes after CBTi. We also hypothesized that the discrepancy between objective and subjective measures of sleep (i.e., sleep-state misperception) would be significantly reduced after CBTi.

## MATERIALS AND METHODS

### Participants

Participants with chronic insomnia were recruited through online and print advertisements posted in the community and from physician referral. Prospective participants were initially screened over the phone for inclusion and exclusion criteria, followed by a semi-structured in-person interview. Potentially eligible participants subsequently underwent a screening polysomnographic (PSG) recording to rule out the presence of other sleep disorders contributing to insomnia symptoms.

Participants meeting the 3^rd^ version of the International Classification of Sleep Disorders (ICSD-3) diagnostic criteria for chronic insomnia disorder for at least 6 months were included in the study^1^. The ICSD-3 criteria for chronic insomnia are defined as self-reported difficulties initiating sleep (i.e., sleep onset latency greater than 30 min), difficulties maintaining sleep (i.e., wake after sleep onset (WASO) greater than 30 min), and/or early morning awakenings (i.e., final awakening time earlier than desired by at least 30 min), for at least 3 times a week and for more than 3 months, combined with complaints of daytime functioning.

Exclusion criteria were as follows: being less than 18 years old, a psychiatric condition other than depression and anxiety, medical conditions likely to affect sleep (e.g., epilepsy, multiple sclerosis, Parkinson’s disease, chronic pain, active cancer), other sleep disorders (e.g., moderate to severe sleep apnea defined by apnea-hypopnea index (AHI) > 15/h, restless legs syndrome, periodic limb movement disorder defined by an index during sleep > 15/h), a major cardiovascular condition or intervention, a recent severe infection, poor cognitive function (defined by a diagnosis of dementia or a Montreal Cognitive Assessment – MoCA – score <26^37^), shift work or changes in time zones over the past 2 months, and monthly use of recreational drugs or weekly use prescription drugs that might affect sleep. All participants signed a written informed consent form before entering the study, which was approved by the Concordia University Human Research Ethics Committee. A total of sixty-two participants out of 354 were found eligible for the study and randomized to one of the two groups (see below). Their demographic information is presented in **Table 1**.

**Table 1 -.**
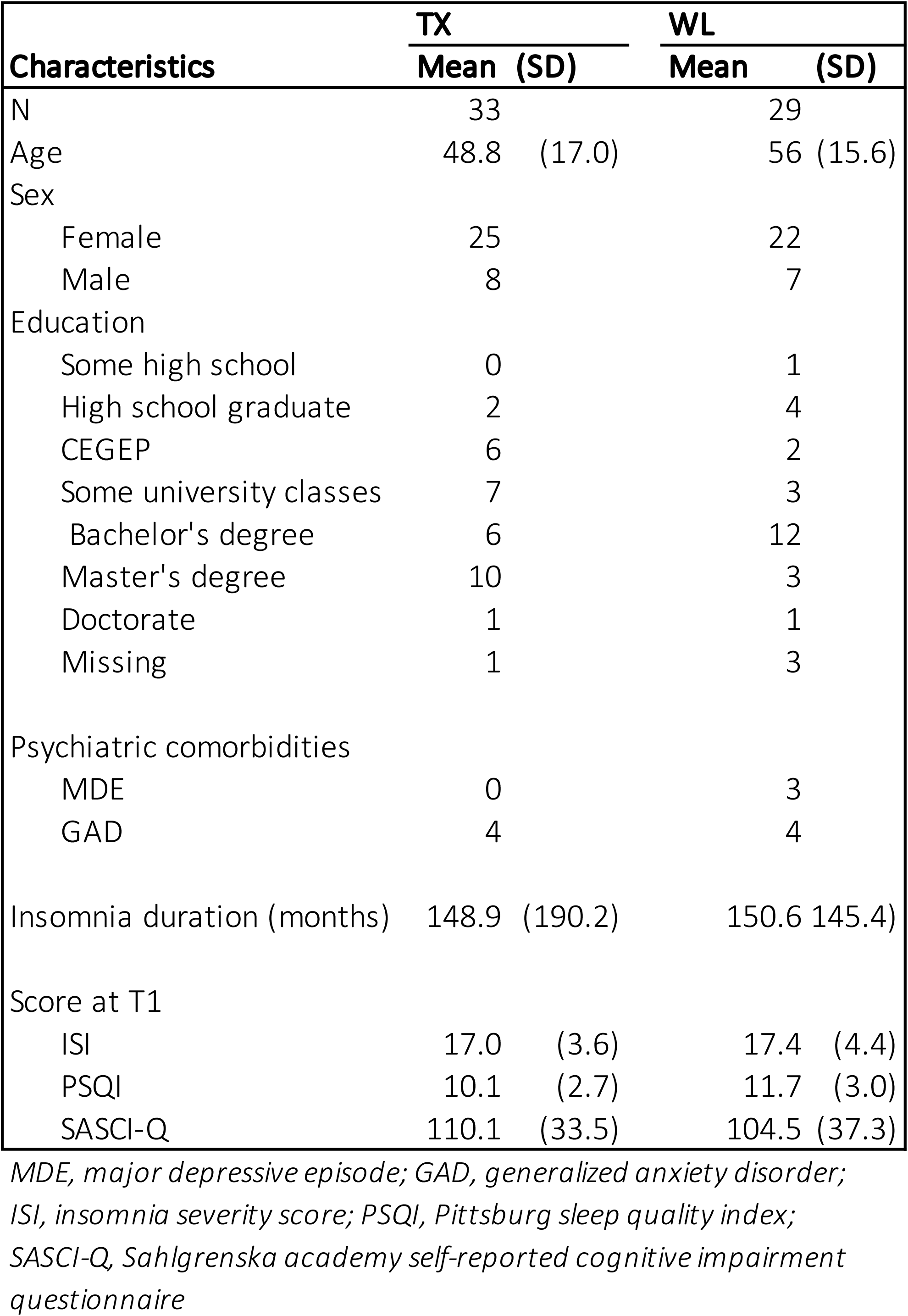
Demographics.

| Characteristics | TX |  | WL |  |
| --- | --- | --- | --- | --- |
|  | Mean | (SD) | Mean | (SD) |
| N | 33 |  | 29 |  |
| Age | 48.8 | (17.0) | 56 | (15.6) |
| Sex |  |  |  |  |
| Female | 25 |  | 22 |  |
| Male | 8 |  | 7 |  |
| Education |  |  |  |  |
| Some high school | 0 |  | 1 |  |
| High school graduate | 2 |  | 4 |  |
| CEGEP | 6 |  | 2 |  |
| Some university classes | 7 |  | 3 |  |
| Bachelor's degree | 6 |  | 12 |  |
| Master's degree | 10 |  | 3 |  |
| Doctorate | 1 |  | 1 |  |
| Missing | 1 |  | 3 |  |
| Psychiatric comorbidities |  |  |  |  |
| MDE | 0 |  | 3 |  |
| GAD | 4 |  | 4 |  |
| Insomnia duration (months) | 148.9 | (190.2) | 150.6 | 145.4) |
| Score at T1 |  |  |  |  |
| ISI | 17.0 | (3.6) | 17.4 | (4.4) |
| PSQI | 10.1 | (2.7) | 11.7 | (3.0) |
| SASCI-Q | 110.1 | (33.5) | 104.5 | (37.3) |

*MDE, major depressive episode; GAD, generalized anxiety disorder;*
*ISI, insomnia severity score; PSQI, Pittsburg sleep quality index;*
*SASCI-Q, Sahlgrenska academy self-reported cognitive impairment questionnaire*

### Protocol

Eligible participants were enrolled in a randomized controlled trial comprised of an immediate CBTi arm and a wait-list control arm (see **Figure 1A** for the study design). Within a month after the initial PSG screening, participants completed a baseline assessment (T1). Participants completed a sleep diary for two weeks before coming back to the lab for another PSG recording. In addition to the PSG, participants filled out questionnaires in the evening and performed cognitive tasks (i.e., ANT and N-back) the next morning. Following this baseline assessment, participants were randomized to either a 3-month CBTi program (8 group sessions; TX group) or a 3-month wait-list period (WL group) stratified by age and sex, with a 1:1 ratio. Thirty-three participants were randomized to the TX group and 29 participants were randomized to the WL group. Three months post-randomization (T2), participants from both groups came back to the laboratory and underwent the same protocol as the baseline assessment (PSG, cognitive tasks, questionnaires) followed by a 2-week sleep diary. Following T2 assessment, participants from the WL group received a 3-month CBTi program. At 6-month post-randomization (T3), participants from the WL group came back to the laboratory where they underwent the same protocol.

**Figure 1.**
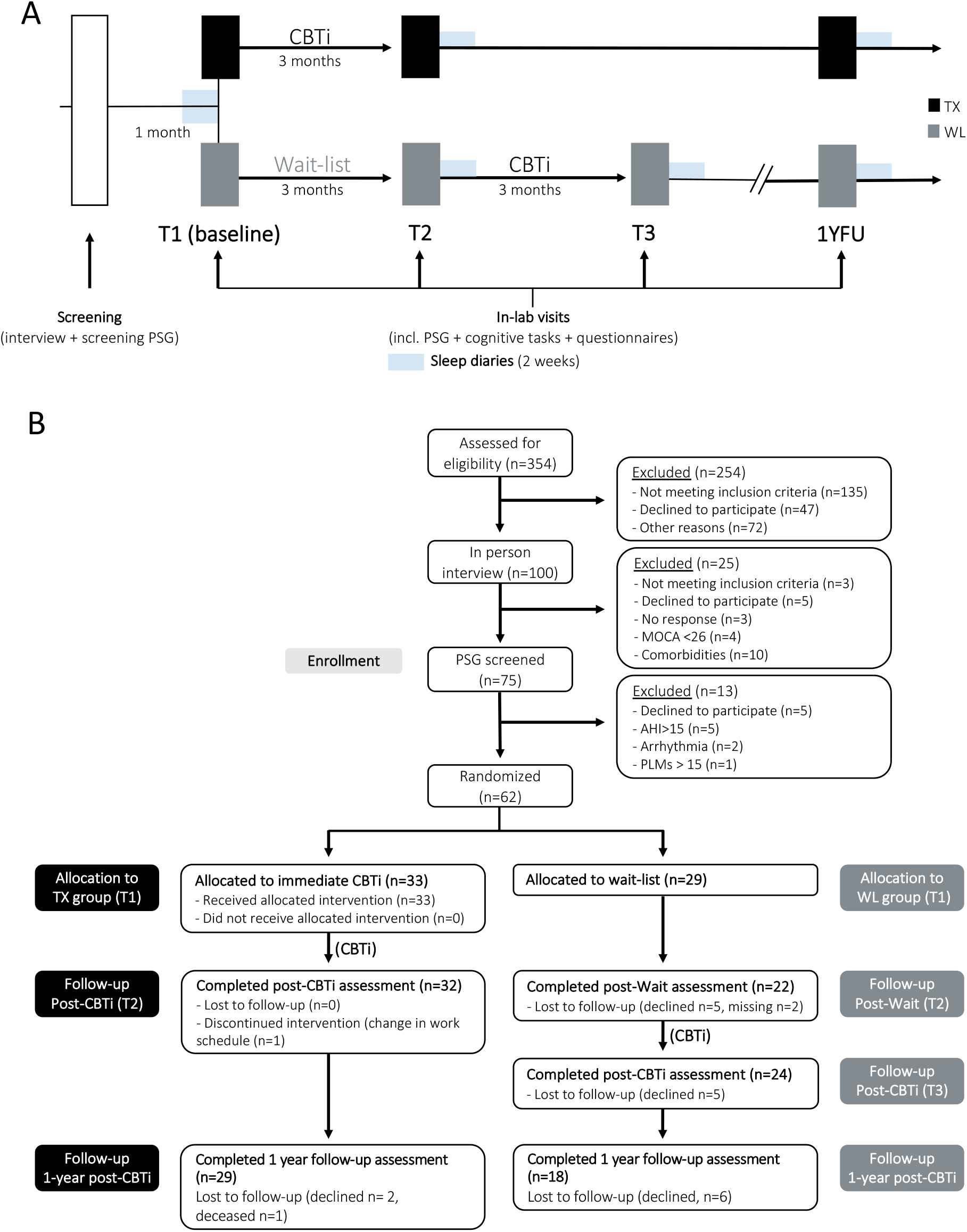
Study design and participant flow. (A) Participants underwent a screening, comprised of a clinical interview and polysomnography (PSG) to screen for inclusion and exclusion criteria. A the first timepoint (T1) participants underwent a baseline assessment: PSG recording, filled out questionnaires, completed computerized cognitive tasks the next morning, sleep diaries for 2 weeks, and were randomized into either the immediate cognitive-behavioral therapy for insomnia group (TX group - black) or the wait-list group (WL group - grey). After 8 weeks (T2), participants underwent the same assessments as baseline. Participants in the WL group received a CBTi and were invited to come in the lab for an additional assessment (T3) similar to T1 and T2. Finally, all participants were invited to repeat the same assessments one year after completion of CBTi (1YFU) (B) Consort flow chart of the participants PSG = polysomnography, AHI = Apnea-Hypopnea Index, MoCA = Montreal Cognitive Assessment, PLMs = Periodic Limb Movements during sleep, CBTi = Cognitive-Behavioral Therapy for insomnia.

One year after the completion of the CBTi program (1YFU assessment), participants from both groups were contacted to undergo the same protocol (PSG, cognitive tasks, questionnaires, 2-week sleep diary) (see **Figure 1B** for depiction of the CONSORT flow chart). This study was registered as a clinical trial (ISRCTN13983243, https://doi.org/10.1186/ISRCTN13983243).

### CBTi intervention

Participants in the TX group (after T1) and in the WL group (after T2) underwent an 8-session group CBTi within 3 months, which consisted of psychoeducation about sleep and circadian rhythms, stimulus control, sleep restriction, sleep hygiene, cognitive therapy, and relaxation based on Morin & Espie^38^. Each group included 2 to 6 participants, for a total of 14 CBTi groups in the present study. The treatment was conducted by a trained psychologist. Each weekly session lasted 90 minutes.

### Measures

#### Questionnaires

At each time point (T1, T2, T3, 1YFU), participants completed the following questionnaires:

##### Insomnia Severity Index (ISI)

The ISI is a widely used self-report tool for assessing the nature, severity, and impact of current insomnia symptoms^20,39^. It consists of 7 Likert-scale questions with a total score ranging from 0 to 28 (with higher scores indicating more severe insomnia) and its overall Cronbach’s α was .68. The change in ISI score was our primary outcome for this study. Individuals who reported a decrease in ISI by 7 points or more following CBTi were considered to be Responders, while those who did not were considered Non-Responders^40^. Those who had an ISI score below 8 after CBTi were considered to be Remitters^20^.

##### Pittsburgh Sleep Quality Index (PSQI)

The PSQI is a self-report measure of general sleep quality over the past month^21^. It consists of 7 sub-components (subjective sleep quality, sleep latency, sleep duration, habitual sleep efficiency, sleep disturbances, use of medication, and daytime dysfunction) calculated through 4 questions on the timing of sleep behaviors and 6 Likert-scale questions on sleep habits. Total PSQI score ranges from 0 to 21 (with higher scores indicating worse sleep quality) and its overall Cronbach’s α was .44.

##### Sahlgrenska Academy Self-reported Cognitive Impairment Questionnaire (SASCI-Q)

The SASCI-Q is a 29-item questionnaire assessing current self-reported cognitive functioning and cognitive decline, spanning different cognitive domains such as memory, attention, executive function, language and mixed domains^41^. Total SASCI-Q scores range from 29 to 203, with a higher score indicating more cognitive impairment, and its overall Cronbach’s α was .96.

#### Sleep diaries

At each time point (T1, T2, T3, 1YFU), participants provided information about their sleep using the Consensus sleep diary^42^ every day during 2 weeks. They reported their bedtime, sleep onset latency, number of nocturnal awakenings, wake after sleep onset, wake-up time, and out of bed time. They also estimated their sleep duration and time spent awake as well as their sleep satisfaction rating using a 5-point rating scale (from 1: very bad sleep to 5: very good sleep). For each sleep variable, measures were averaged over 2 weeks at each time point. Only sleep diary data with more than 5 days completed per timepoint were included in the analyses.

#### Polysomnographic (PSG) recording

Whole-night PSG recording was used for all in-lab nights. PSG included EEG, EOG, EMG, ECG. The 17 scalp electrodes (F7, F3, Fz, F4, F8, T3, C3, Cz, C4, T4, P3, Pz, P4, O1, O2, M1, M2) were placed according to the international 10-20 system. During the screening PSG (first night), periodic leg movements (legs EMG) and Apnea-Hypopnea Index (thermistance, nasal canula, thoraco-abdominal belts and oximeter) were also computed in order to exclude other sleep disorders. The EEG signal was recorded with a Somnomedics amplifier (Somnomedics GmbH, Germany), sampled at 512 Hz. EEG recordings were referenced online to Pz, and for the offline analyses the EEG signals were re-referenced to the contra-lateral mastoids (M1, M2). All sleep scoring and analyses were conducted using the Wonambi python toolbox (https://wonambi-python.github.io). For each recording, at least two scorers determined the different sleep stages (NREM 1, 2, 3, REM sleep and wake) according to the AASM rules for each recorded night of sleep^43^. From the scoring of the sleep architecture, we computed sleep onset latency (SOL), wake after sleep onset (WASO), sleep duration (total sleep time; TST), the percentage of each sleep stage relative to the total sleep period (TSP; from sleep onset to wake up time) and sleep efficiency (TST/time in bed*100).

#### Sleep-state misperception measures

In the morning following each overnight PSG study, participants reported their subjective estimation of time to fall asleep (sleep latency) and sleep duration in a night review (NR) questionnaire. Measures of sleep-state misperception (SSM) were extracted by subtracting subjective estimates from objective (PSG) assessments and included SSM-TST (objective TST minus subjective sleep duration) and SSM-SOL (objective SOL minus subjective sleep latency)^27^. While SSM-Wake (objective WASO minus subjective wake duration) is traditionally computed in studies on sleep misperception, we were no able to assess subjective Wake duration in the night review.

#### Cognitive tasks

The morning after each night at the lab, objective cognitive function was assessed using two tasks running on a computer with Inquisit 4.0.9.0 software (Millisecond, Seattle, WA, https://www.millisecond.com/), assessing cognitive domains that were found impaired in insomnia, i.e., working memory and attention^44^.

The Attention Network Task (ANT) was used to test the three networks of attention (i.e., alerting, orienting and executive function (conflict resolution efficiency)^45^ - see **Figure S1A** for the design). Participants had to respond to the direction of the center arrow (target) of an array of 5 arrows (flanker arrows) for a total of 144 trials. The array of arrows appeared on either side of the screen and were cued by a flashing box. Cues were presented 0-800 ms (mean 400 ms) prior to the presentation of the arrows, highlighting the location of the array of arrows and were of three different types: no cue, double cue, or single cue which could validly be cueing the location of the array of arrows or not. The direction of the flanker arrows was either congruent or incongruent with the center (target) arrow. Efficiency of the alerting network corresponds to vigilance and is measured by the ability to use a cue to probe attention (RT_no cue_ – RT_double cue_). Efficiency of the orienting network is measured by the ability to select and respond to a specific stimulus (RT_invalid cue_ – RT_valid cue_). The executive component corresponds to the ability to resolve conflict between incongruent and congruent information (RT_incongruent flanker_ – RT_congruent flanker_). The following variables were extracted: % Accuracy, Reaction time (RT), and reaction time difference between stimuli, for each of the 3 attention networks (alerting, orienting and executive function).

The second task was the N-back task, which tests working memory performance^46,47^ (see Figure S1B for the design). The task consisted in the presentation of a series of letters at a fixed frequency (2500 ms), and participants were asked to press a button each time the current stimulus was identical to the stimulus presented N-letters before. We assessed three different difficulty levels: 1-, 2- and 3-back, each level comprising 45 trials (including 15 target trials) randomly presented for a total of 135 trials. Accuracy and reaction times were extracted across all difficulty levels, as well as for each difficulty level separately.

### Statistical analyses

Statistical analyses were performed using RStudio 1.2.50 (RStudio, Inc., Boston, MA).

An Intent-to-Treat analysis was conducted on our primary outcome measure (i.e., ISI score) using mixed-model analysis of variance (ANOVA) with Group as a between-subject factor (TX group vs WL group) and Time as a within-subject factor (T1 vs T2) with imputation on the 62 individuals with chronic insomnia randomized into the study to investigate the effect of CBTi. In addition, we performed a hierarchical linear modeling (HLM) analysis to evaluate changes in insomnia severity (ISI score) by pooling data from both groups from pre (T1)- to post-CBTi (T2 for TX and T3 for WL) and one-year follow-up (1YFU). Given the variability in attrition depending on the measures (see Attrition of each measure in **Supplementary Methods**), we decided to conduct per protocol analyses to investigate the efficacy of CBTi on our secondary outcomes, including subjective perception of sleep (i.e., sleep variables from sleep diaries), objective sleep architecture (i.e., sleep variables from PSG), sleep-state misperception (SSM-TST and SSM-SOL), subjective perception of cognitive abilities (SASCI-Q score) and objective performance (accuracy and reaction times) at attentional (ANT) and working memory (N-back) tasks. Mixed-model analyses of variance (ANOVA) with Group as a between-subject factor (TX group vs WL group) and Time (T1 vs T2) and Group*Time interaction were used to investigate the effects of CBTi compared to a wait-list period. To increase statistical power and investigate the effects of CBTi, we also performed complementary analyses on these secondary outcomes by pooling both groups Post-CBTi (T2 for TX group and T3 for WL group). Mixed-model analyses of variance (ANOVA) with Intervention as a within-subject factor (Pre-CBTi(T1) vs Post-CBTi) were used on measures with complete data. Participants were asked to complete the ISI at the one-year follow-up and were given the option of completing an overnight PSG with all the other measurements. Many participants elected not to complete this portion of the study. Given the large attrition for most secondary outcomes, one-year follow-up data for secondary variables were not included in the final analyses (see Attrition of each measure in **Supplementary Methods**).

All analyses were conducted while adjusting for age and sex. Level of education was included as an additional covariate in the analyses on cognitive performances. Normality of distribution was checked with Shapiro tests and homogeneity of variance was tested with Levene tests. Only if variance was not homogeneous, we used non-parametric tests (Friedmann Test or Wald-Type Statistic) and when tests did not report the same results, both analyses were reported, otherwise only ANOVAs were reported. Degrees of freedom were corrected according to the Greenhouse–Geisser method, when appropriate. Within-group effect sizes were calculated to indicate the degree of change in response to Time/Intervention using Hedges’s *g* (corrects for small sample size). The level of significance was set to a p-value of <.05 and p-values were adjusted for multiple comparisons (Benjamini-Hochberg/FDR correction). For significant results, both raw (*P*) and adjusted p-values (*Q*) were reported when necessary. Exploratory bivariate Spearman’s Rho correlations were conducted between primary (ISI score) and secondary measures in both groups to investigate the relationship between change in insomnia severity and changes in subjective and objective sleep and cognitive abilities.

## RESULTS

Sixty-two individuals with chronic insomnia were randomized to either the immediate CBTi arm (TX group - N=33) or a wait period (WL group - N=29). Participants were middle-aged (52 ± 16 years old), in majority female (75.8%) and mostly (69%) holders of university diploma (see **Table 1** for demographics).

At 3-months post-randomization (T2), 32 participants (96.9%) from the TX group and 22 participants (75.8%) from the WL group filled the ISI questionnaire (primary outcome). We found no difference in age (*t*-test *P* >.05) or sex (Fisher Exact Test *P* >.05) between participants who did or did not provide ISI score at T2 in the TX group (P > .05) but there was a significant difference in age in the WL group, with participants who did not provide ISI score at T2 being younger (38.5 ± 21 years old) than the participants who did provide measures (56.5 ± 10 years old; t= 17.7, P <.001). For the pooled data Post-CBTi, out of the 62 participants who filled the ISI Pre-CBTi (T1), 56 (90.3%) filled the ISI after completion of the CBTi program. There were no differences in gender or age between participants who did or did not provide measures Post-CBTi (P > .05). Attrition rates of the secondary measures can be found in the **Supplemental Methods**.

### CBTi reduces insomnia severity

At 3-months post-randomization (T1 versus T2), we found a Group*Time interaction (*F*(1,110) = 17.71; *P* <.001) with main effects of Time (*F*(1,110) = 32.82; *P* <.001) and Group (*F*(1,102) = 16.79; *P* <.001) due to the large reduction in ISI score only in the TX group at T2 (*P* <.001; *g’* = 1.66; see Table 2 and Figure 2A). On average, the TX group reduced their ISI score by 7.5 ± 4.88 points after CBTi while the WL group did not show a significant change (−0.98 ± 4.30 points) in ISI score. In the TX group, 16 (50%) out of 32 participants were considered as responders (ISI decreasing by at least 8 points) and 12 (37.5%) were considered in remission (ISI score < 8) at T2, while only one participant (4.54%) from the WL group was in remission at T2.

**Table 2 -.**
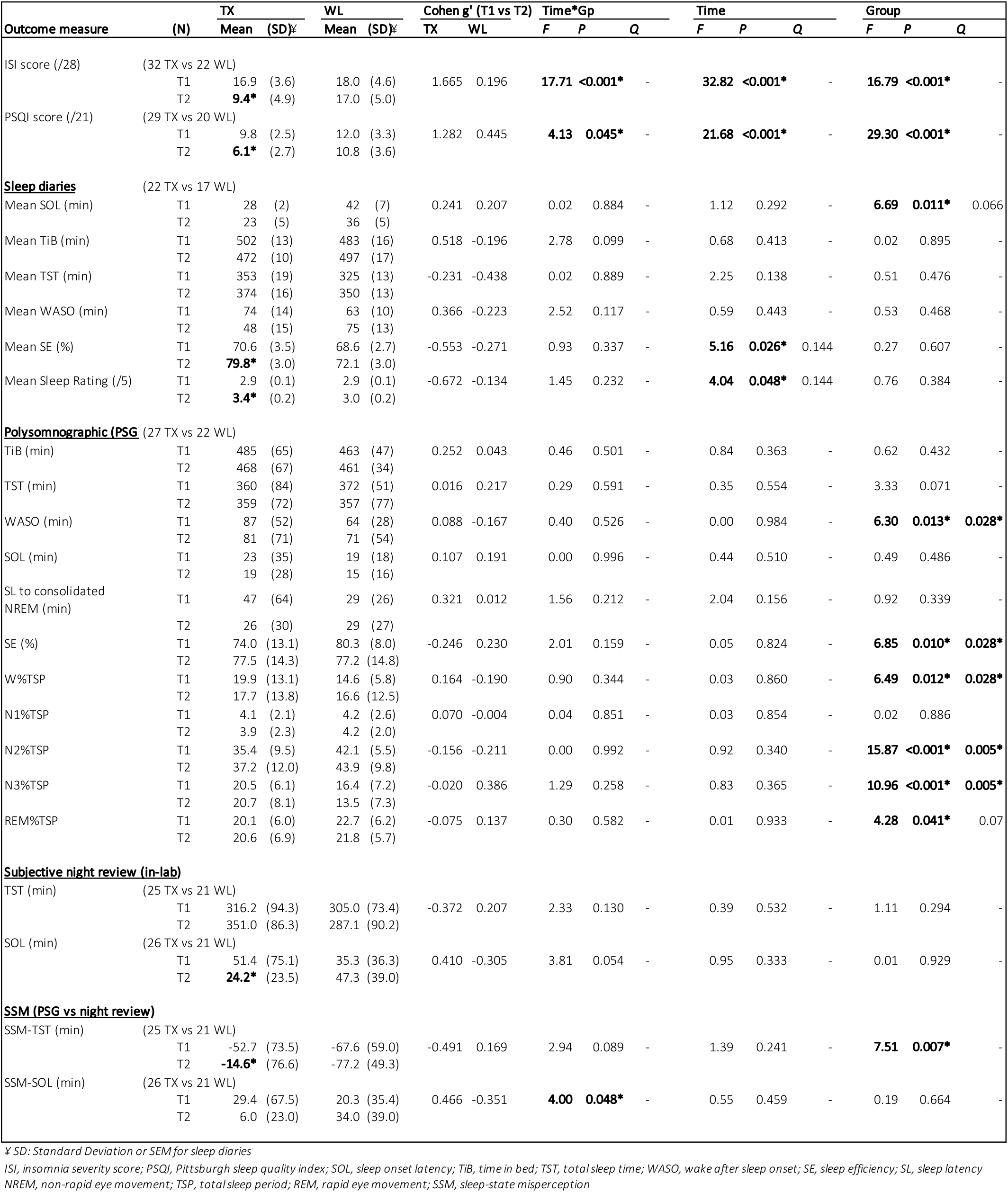
Sleep measures (T1 versus T2)

**Figure 2.**
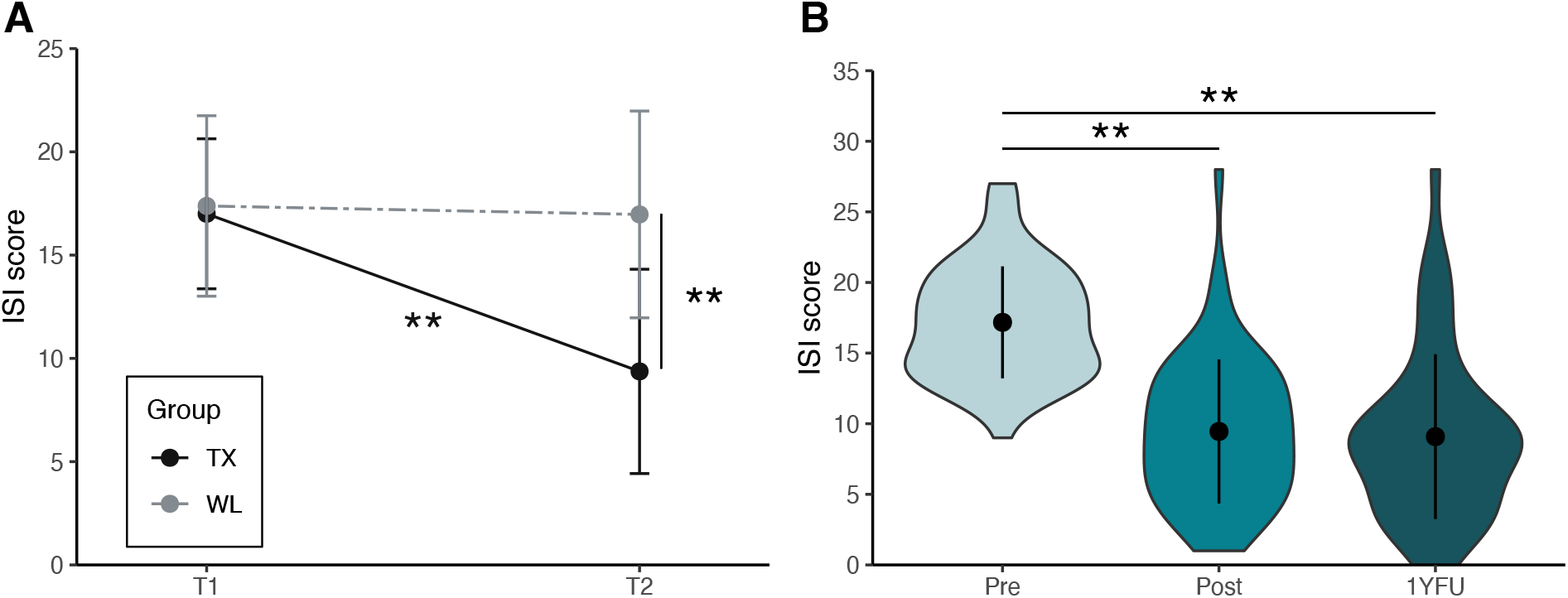
Insomnia Severity – Effect of CBTi on ISI score. (A) Mean ISI score (± SD) at baseline (T1) and 3 months post-randomization (T2) for the TX group (black) and the WL group (grey). (B) Mean ISI score (± SD) across all participants (pooled groups) at Pre- (T1), Post-CBTi (T2 for TX and T3 for WL) and 1 year after completion of the CBTi (1YFU). Asterisks represent significance (P): ** <0.001

Complementary analyses on pooled data Post-CBTi (T2 for TX group and T3 for WL group; N=56) from both groups revealed similar large reduction in ISI score with a main effect of Intervention (*F*(1,108) = 88.4; *P* <.001; *g’* = 1.69) and similar response and remission rates. Indeed, out of 56 participants who filled the ISI questionnaire after CBTi, 30 were considered as responders (53.5%) and 22 were in remission (39.2%). HLM analyses on the trajectory of ISI score combining post-CBTi assessments (i.e. T2 for TX and T3 for WL as well as 1YFU) for both groups revealed a similar decrease in ISI score Post-CBTi (Pre-CBTi vs Post-CBTi, *t*=12.33, *P* <.001, *g*’ = 1.76, *beta* =-7.76) that was maintained one-year after completion of the intervention (Pre-CBTi vs 1YFU: *t*=12.6, *P* <.001, *g’*= 1.71, *beta* =-8.2 and Post-CBTi vs 1YFU: *t*=.61, *P* =.816, *g’* =.053; see **Table S1 and Figure 2B**). Out of the 47 participants who filled out the ISI at 1YFU, 18 (38.2%) were in remission a year after CBTi.

### CBTi improves self-reported sleep quality

At 3-months post-randomization (T1 versus T2), per-protocol analyses showed a Group*Time interaction for PSQI score (*F*(1,86) = 4.13; *P* =.045), due to a significant decrease in PSQI score in the TX group only (TX:-3.8 ± 0.77pt, *P* <.001, N=29; *g’* = -1.28; WL:-1.3 ± 0.93pt, *P* =.16, N=20; *g’* = -0.44; see **Table 2**). Self-reported assessment of sleep (e.g., duration, latency, efficiency, satisfaction) using sleep diaries (TX N=20, WL N=17) did not reveal any significant interaction, however, we found an effect of Time for subjective sleep efficiency (SE; *F*(1,72) = 5.1; *P* =.02, *Q* =.14) and subjective sleep satisfaction (*F*(1,72) = 4.04; *P* =.04, *Q* =.14) due to a large increase in both measures in the TX group only (see **Table 2** and **Figure 3A**). However, note that these effects did not survive multiple comparisons.

**Figure 3.**
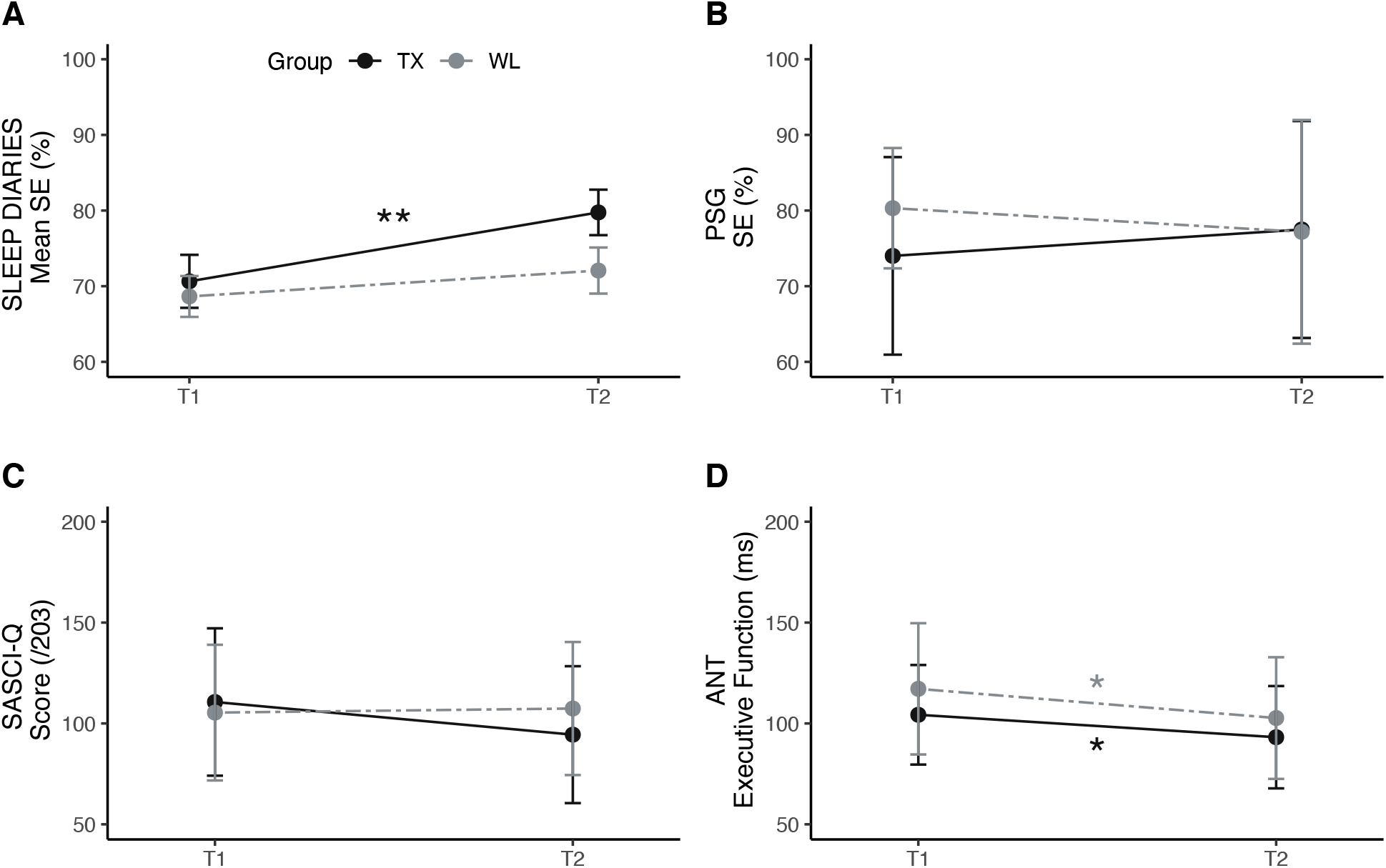
Multimodal assessment of CBTi efficacy on sleep and cognition. (A) Mean sleep efficiency (± SEM) extracted from 14-day sleep diaries at baseline (T1) and 3 months post-randomization (T2) for TX group (black; N=20) and WL group (grey; N=17). (B) Mean sleep efficiency (± SD) extracted from polysomnographic recording at baseline (T1) and 3 months post-randomization (T2) for TX group (N=27) and WL group (N=22). (C) Mean Sahlgrenska Academy Self-reported Cognitive impairment questionnaire (SASCI-Q) (± SD) at baseline (T1) and 3 months post-randomization (T2) for TX group (N=26) and WL group (N=21). (D) Mean reaction time (± SEM) extracted from the executive function component of the Attention Network Test (ANT) at baseline (T1) and 3 months post-randomization (T2) for TX group (N=26) and WL group (N=21). Asterisks represent significance (P) of post-hoc comparisons adjusted for Age and Sex (and education level for C & D): ** <0.001, * <0.05

Complementary analyses on pooled data Post-CBTi from both groups revealed similar changes in subjective sleep quality, including the reduction in PSQI score (−3.75 ± 0.59 point, *P* <.001, *g’* = -1.27, N=54; see **Table S2**) as well as better sleep diaries-determined SE (+8.85 ± 2.82 point; *P* =.002; *Q* =.012, *g*’ =-0.62, N=37) and sleep satisfaction (+0.43 ± 2.82 points; *P* =.015; *Q* =.034, *g’* = -0.57, N=37; see **Table S2**). Moreover, we found a decreased self-reported SOL (*F*(1,70) = 5.9; *P* =.017; *Q* =.034) by 11 ± 4min on average (*g’* =0.54) and wake duration (WASO; *F*(1,70) = 4.8; *P* =.031; *Q* =.038) by 26 ± 6min on average (*g*’ =0.43). They also reported a decreased time spent in bed (TiB; *F*(1,70) = 4.7; *P* =.032; *Q* =.038) by 28 ± 7min on average (*g’* =0.46), but there was no change in self-reported total sleep duration (*P* >.05; see **Table S2**).

### CBTi does not improve objective sleep measures

At 3-months post-randomization (TX N=27, WL N=22), per-protocol investigation of sleep architecture using PSG recording (e.g., sleep efficiency, total sleep time, sleep latency, total time spent awake after sleep onset) did not reveal any Group*Time interaction (all *P* >.05; see **Table 2** and **Figure 3B**). Complementary analyses on pooled data Post-CBTi (N=46) revealed similar lack of change in objective sleep measures (all *P* >.05; see **Table S2**).

### Degree of discrepancy between objective and subjective sleep decreases after CBTi

At 3-months post-randomization, there was no Group*Time interaction for both subjective (Night Review) and objective (PSG) in-lab measures of sleep duration (TST) and sleep onset latency (SOL; see **Table 2**). However, per-protocol analyses showed a Group*Time interaction (*F*(1,87) = 4; *P* =.048) in the misperception of sleep onset latency (SSM-SOL) driven by a trend to decrease SSM-SOL in the TX group only (*P* =.062; *g’* =0.46; see **Table 2**). Direct comparison of subjective SOL with objective SOL with pairwise *t*-test revealed significant difference at T1 in both groups (TX: *t* = -2.51, *P* =.01, N=26; WL: *t* = - 2.88; *P* =.009, N=21) confirming the presence of SOL misperception. However, at T2, while the WL group still exhibited a significant discrepancy between objective and subjective SOL (*t* = -3.94; *P* <.001), the TX group no longer showed SSM-SOL (*t* = -1.07; *P* =.29).

Concerning change in the misperception of sleep duration (SSM-TST), while the Group*Time interaction was only marginally significant (*F*(1,86) = 2.94; *P* =.089), we found a main effect of Group (*F*(1,86) = 7.51, *P* =.007). The significant difference of SSM-TST at T2 (*P* =.002) was driven by a decrease in the degree of discrepancy in SSM-TST in TX group only (*P* =.045; *g’* =0.49; see **Table 2 and Figure 4A**). Indeed, when we directly compared subjective TST with objective TST using a pairwise *t*-test, we found that at T1 both groups showed significant differences between subjective and objective TST (all *P* <.001), whereas at T2, the WL group continued to exhibit a significant difference between objective and subjective TST (*t* = 7.15; *P* <.001), but the TX group no longer showed any difference (*t* = 1.04; *P* =.30 – see **Figure 4B**).

**Figure 4.**
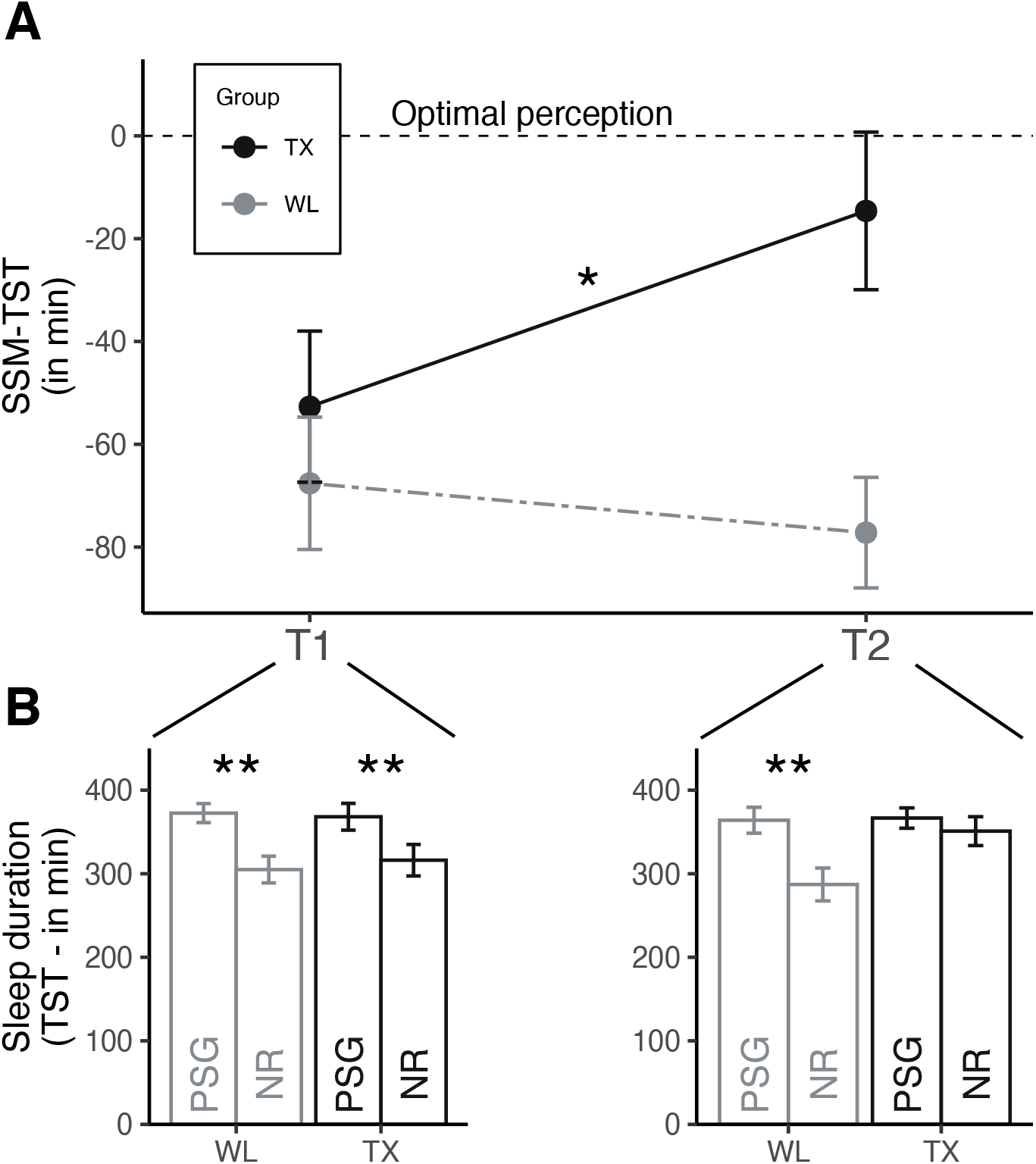
CBTi efficacy on sleep-state misperception of sleep duration (SSM-TST) (A) Mean sleep-state misperception on sleep duration (± SD) computed from subjective (night review (NR)) minus objective (polysomnographic recording (PSG)) sleep duration at baseline (T1) and 3 months post-randomization (T2) for TX group (black; N=26) and WL group (grey; N=21). (B) Mean sleep duration (± SD) extracted from PSG recording and NR at baseline (T1 – left panel) and 3 months post-randomization (T2 – right panel) for TX and WL groups. Asterisks represent significance (P): ** <0.001, * <0.05

Complementary analyses on pooled data Post-CBTi (N=46) revealed similar reduction in SSM-SOL and SSM-TST after the Intervention (see **Table S2**). Indeed, while there were significant differences between objective and subjective measure of TST and SOL Pre-CBTi (all *P* <.001), both SSM-SOL (*t* = - 1.78; *P* =.08) and SSM-TST (*t* = 1.47; *P* =.14) were no longer significantly different Post-CBTi.

### CBTi does not improve self-reported cognitive functioning

At 3-months post-randomization (TX N=27, WL N=20), we found no Group*Time interaction on the SASCI questionnaire (SASCI-Q) score (*F*(1,87) = 1.6; *P* =.21; see **Table 3** and **Figure 3C**) suggesting no change in subjective cognitive functioning after CBTi. Complementary analyses on pooled data Post-CBTi (N=43) revealed a trend for SASCI-Q score reduction Post-CBTi (*P* =.079, -12.7 ± 7.5pt, *g’* = -0.38, see **Table S3**).

**Table 3 -.** Cognitive measures (T1 vs T2)

| Outcome measure | (N) | TX |  | WL |  | Cohen g' (T1 vs T2) |  | Time*Gp |  |  | Time |  |  | Group |  |  |
| --- | --- | --- | --- | --- | --- | --- | --- | --- | --- | --- | --- | --- | --- | --- | --- | --- |
|  |  | Mean | (SD) | Mean | (SD) | TX | WL | F | P | Q | F | P | Q | F | P | Q |
| **Subjective cognitive functioning** (27 TX vs 20 WL) |  |  |  |  |  |  |  |  |  |  |  |  |  |  |  |  |
| SASCI-Q score (/203) | T1 | 109 | (36.5) | 101 | (33.6) | 0.470 | -0.058 | 1.60 | 0.209 | - | 1.41 | 0.2385 | - | 0.01 | 0.907 | - |
|  | T2 | 93 | (33.9) | 103 | (33.0) |  |  |  |  |  |  |  |  |  |  |  |
| **Objective cognitive functioning** |  |  |  |  |  |  |  |  |  |  |  |  |  |  |  |  |
| **ANT** (26 TX vs 21 WL) |  |  |  |  |  |  |  |  |  |  |  |  |  |  |  |  |
| Accuracy (% correct) | T1 | 86.1 | (19.9) | 87.9 | (21.5) | -0.410 | -0.469 | 0.02 | 0.888 | - | **4.48** | **0.037\*** | 0.092 | 0.74 | 0.393 | - |
|  | T2 | **90.7\*** | (10.1) | **93.1\*** | (9.0) |  |  |  |  |  |  |  |  |  |  |  |
| Reaction time - correct (ms) | T1 | 665 | (94.7) | 666 | (55.0) | 0.221 | 0.028 | 0.16 | 0.702 | - | 0.26 | 0.622 | - | **5.50** | **0.020\*** | 0.1 |
|  | T2 | 650 | (102.6) | 664 | (60.3) |  |  |  |  |  |  |  |  |  |  |  |
| Alerting (ms) | T1 | 25 | (27.3) | 22 | (33.8) | -0.152 | -0.280 | 0.19 | 0.660 | - | 1.19 | 0.2785 | - | 0.00 | 0.958 | - |
|  | T2 | 29 | (26.9) | 31 | (32.7) |  |  |  |  |  |  |  |  |  |  |  |
| Orienting (ms) | T1 | 86 | (51.4) | 91 | (43.7) | -0.185 | -0.031 | 0.14 | 0.713 | - | 0.32 | 0.5748 | - | 0.06 | 0.803 | - |
|  | T2 | 93 | (44.2) | 92 | (28.6) |  |  |  |  |  |  |  |  |  |  |  |
| Executive function (ms) | T1 | 108 | (24.7) | 120 | (32.5) | 0.401 | 0.523 | 0.09 | 0.771 | - | **4.88** | **0.029\*** | 0.092 | 3.03 | 0.086 | - |
|  | T2 | **97\*** | (25.4) | **106\*** | (30.2) |  |  |  |  |  |  |  |  |  |  |  |
| **N-back** (24 TX vs 18 WL) |  |  |  |  |  |  |  |  |  |  |  |  |  |  |  |  |
| Accuracy (% correct) | T1 | 86.6 | (7.5) | 91.0 | (5.0) | -0.456 | -0.078 | 0.73 | 0.395 | - | 1.81 | 0.1822 | - | **8.50** | **0.005\*** | **0.026\*** |
|  | T2 | 89.1 | (6.5) | 91.4 | (5.7) |  |  |  |  |  |  |  |  |  |  |  |
| Accuracy - 1-back (%) | T1 | 94.0 | (8.0) | 98.2 | (3.7) | -0.504 | 0.000 | 1.31 | 0.256 | - | 1.74 | 0.1907 | - | **7.00** | **0.010\*** | **0.026\*** |
|  | T2 | 96.5 | (3.5) | 98.2 | (4.6) |  |  |  |  |  |  |  |  |  |  |  |
| Accuracy - 2-back (%) | T1 | 86.7 | (10.0) | 90.5 | (8.6) | -0.399 | -0.194 | 0.22 | 0.642 | - | 2.03 | 0.1579 | - | 2.03 | 0.158 | - |
|  | T2 | 89.8 | (8.9) | 92.1 | (7.1) |  |  |  |  |  |  |  |  |  |  |  |
| Accuracy - 3-back (%) | T1 | 79.1 | (8.7) | 84.3 | (6.3) | -0.267 | 0.035 | 0.47 | 0.495 | - | 0.40 | 0.5299 | - | **7.01** | **0.010\*** | **0.026\*** |
|  | T2 | 80.9 | (8.7) | 84.1 | (7.9) |  |  |  |  |  |  |  |  |  |  |  |
| Reaction time (ms) | T1 | 704 | (158) | 715 | (109) | 0.176 | 0.222 | 0.01 | 0.917 | - | 0.81 | 0.3724 | - | 0.12 | 0.730 | - |
|  | T2 | 684 | (122) | 691 | (101) |  |  |  |  |  |  |  |  |  |  |  |
| Reaction time - 1-back (ms) | T1 | 593 | (124) | 588 | (109) | 0.047 | -0.062 | 0.06 | 0.806 | - | 0.00 | 0.998 | - | 0.00 | 0.963 | - |
|  | T2 | 588 | (163) | 595 | (101) |  |  |  |  |  |  |  |  |  |  |  |
| Reaction time - 2-back (ms) | T1 | 764 | (211) | 765 | (137) | 0.408 | 0.332 | 0.03 | 0.863 | - | 2.96 | 0.0895 | - | 0.04 | 0.845 | - |
|  | T2 | 703 | (134) | 715 | (165) |  |  |  |  |  |  |  |  |  |  |  |
| Reaction time - 3-back (ms) | T1 | 789 | (221) | 848 | (165) | 0.186 | 0.179 | 0.00 | 0.987 | - | 0.70 | 0.4054 | - | 1.49 | 0.226 | - |
|  | T2 | 749 | (212) | 809 | (152) |  |  |  |  |  |  |  |  |  |  |  |

SASCI-Q, Sahlgrenska academy self-reported cognitive impairment questionnaire; ANT, attention network task;

### CBTi does not improve attention and working memory performance

At 3-months post-randomization, we did not find a Group*Time interaction in any of the objective measures of cognitive functioning in both the ANT (TX N=26, WL N=21) and N-back tasks (TX N=24, WL N=18; see **Table 3** and **Figure 3D**). However, we found an effect of Time which did not survive multiple comparison correction for overall accuracy (*F*(1,87) = 4.48; *P* =.037, *Q* =.092) and reaction time - executive function (*F*(1,87) = 4.88; *P* =.03, *Q* =.092) at the ANT task, with their performance improving over time across both groups. Complementary analyses on pooled data Post-CBTi revealed similar results for both ANT (N=44) and N-back tasks (N=40; see **Table S3)**.

### Reduction in insomnia severity is associated with improvement in self-reported cognitive function after CBTi

Exploratory associations between change in insomnia severity (primary outcome) and changes in sleep and cognition measures (secondary outcomes) revealed a significant association between the improvement in SASCI-Q score and the improvement in ISI score in the TX group only (*r* = .43, *P* = .025, see **Figure 5**). No significant associations were observed between ISI change and performance changes at the ANT and N-back tasks. Likewise, there was no correlation between change in ISI and changes in objective or subjective sleep variables, including changes in SSM-

**Figure 5.**
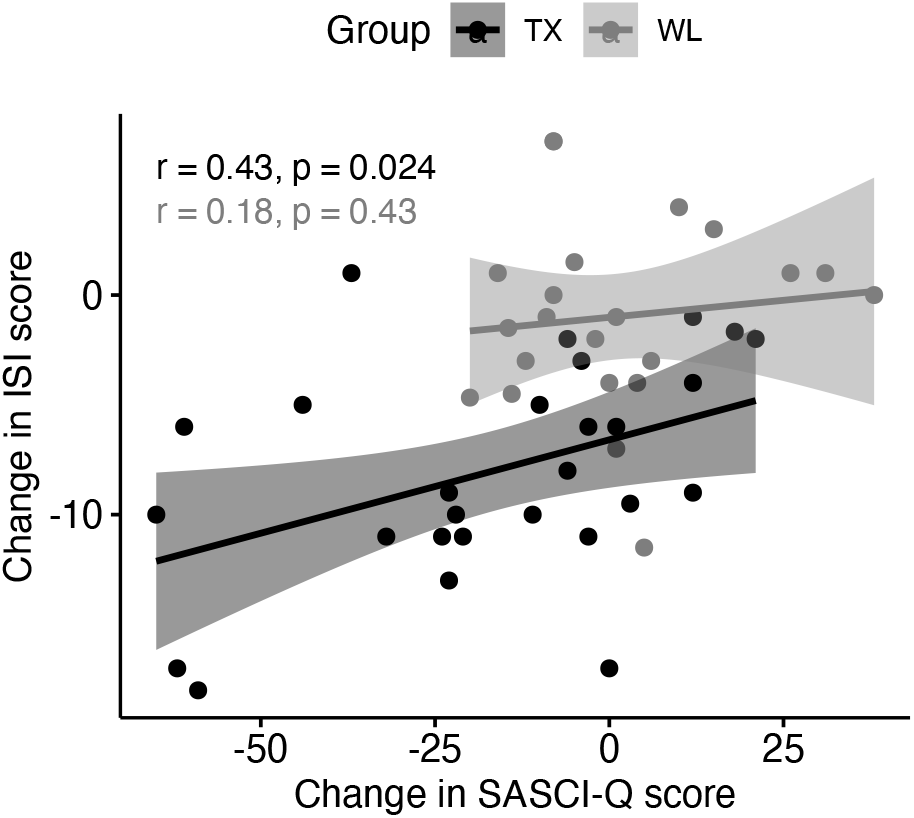
Association between subjective measures of insomnia severity and cognitive abilities. Scatter plot showing correlations between the change in ISI score (T2 minus T1) and the change in Sahlgrenska Academy Self-reported Cognitive impairment questionnaire (SASCI-Q) score (T2 minus T1) in TX group (black) and WL group (grey).

## DISCUSSION

Cognitive-behavioral therapy for insomnia (CBTi) is recommended as the first-line treatment for chronic insomnia. While its efficacy on self-reported insomnia severity has been widely reported, here we used a multimodal approach to specifically assess the efficacy of CBTi on subjective and objective measures of the two main complaints of insomnia, namely sleep difficulties and daytime cognitive impairment. Our analyses revealed a beneficial effect of CBTi on subjective (self-reported) sleep quality/satisfaction while different sleep variables measured objectively did not change following treatment. However, we also reported a reduced sleep-state misperception (i.e., discrepancy between objective and subjective measures) of sleep duration and sleep latency after CBTi. Furthermore, we did not observe any significant improvement in either subjective or objective measures of cognitive function.

Consistent with the extensive literature on the efficacy of CBTi (see Edinger et al., 2021)^12^, we reported a large improvement (*g’*= 1.66-1.69) in insomnia severity (i.e., ISI score) comparable to what has been found in meta-analyses on the efficacy of CBTi delivered in group format for individuals with and without comorbidities^18,24,48^. The beneficial response (>50%) and remission (>35%) rates were also similar to previous trials^12^. The improvement in self-reported sleep quality (i.e., PSQI score) after CBTi is consistent with the improvement of ISI score as sleep satisfaction is considered as the strongest predictor of insomnia severity^20^. Moreover, despite the lack of significant group-by-time interactions on sleep diaries-determined measures, exploratory post-hoc tests also showed an increase in subjective sleep quality (i.e., sleep satisfaction and sleep efficiency) in the TX group but not in the WL group. Furthermore, we saw that complementary analyses on pooled data (Post-CBTi) revealed the significant improvement in self-reported sleep satisfaction/efficiency often reported in the literature ^12^. While participants reported better subjective sleep efficiency, it was not due to a change in self-reported sleep duration but rather to the decrease in time spent in bed (TiB), as expected from the sleep restriction component of CBTi^49^. Restricting the sleep window over several nights is aimed at building sleep pressure, to facilitate sleep onset and promote sleep maintenance at night^50,51^. Accordingly, we observed self-report of shorter sleep onset latency and shorter wake episodes after sleep onset in the pooled analyses with small-to-medium effect sizes (*g’*= 0.24-0.54), in line with previous reports^16,24,48^. On the other hand, the lack of change in self-reported sleep duration after CBTi is consistent with the small to no effects reported in prior studies^16,48^.

While our participants reported less severe subjective symptoms of insomnia, PSG data demonstrated no objective sleep architecture change after CBTi, including sleep latency, time spent awake or sleep efficiency, even when pooling the two groups post-CBTi. The lack of significant improvement in PSG-determined measures of sleep after CBTi has been reported in most studies^12,24^. Indeed, while few studies reported shorter SOL^52,53^, less time spent awake^53,54^, change in time spent in different stages^55^ and change in TST^56^ using PSG, they yielded small effect sizes and results were not consistently replicated across studies^12^. Hence, CBTi seems to primarily improve self-reported sleep measures and thus insomnia symptoms rather than objective measures of sleep quality.

While there is a large number of studies that have investigated the effects of CBTi on subjective and objective measures of sleep separately, there is little data on the how this intervention affects the direct comparison (and discrepancy) between subjective and objective sleep variables, i.e., sleep-state misperception (SSM). SSM has been observed at varying levels in the general population with and without sleep complaints^28,57^. More often reported in individuals with chronic insomnia, such discrepancy between objective and subjective sleep measures led to the consideration of a subtype of chronic insomnia named “paradoxical insomnia”^58^, which no longer exists in the current version of the international classification of sleep disorders^1^. Indeed, SSM appears as a continuum in the sleep characteristics of individuals with chronic insomnia rather than a specific subtype of insomnia^26,28^. In our study, we found at baseline (T1) an overall sleep-state misperception across our participants reflected by a significant mismatch between PSG-determined TST/SOL and self-reported time spent asleep and time to fall asleep. Interestingly, at T2 (3-month post-randomization), our participants in the TX group no longer exhibited significant discrepancy in those measures after CBTi, while those in the WL group still displayed a significant misperception of TST and SOL. Improvement in sleep/wake perception after CBTi was reported in two studies on older adults using actigraphy/sleep diaries^29,30^ but only one study reported a PSG-determined change in sleep latency misperception after CBTi in seniors^31^. Our study extends these findings by additionally demonstrating an improvement in sleep duration (mis)perception after CBTi, in a sample of individuals with chronic insomnia with a larger range of age. Importantly our findings cannot be explained by a mere improvement of self-reported sleep variables in the absence of change in the corresponding PSG variables, as we did not observe significant improvement in either objective or subjective sleep duration. It has recently been demonstrated that, among good sleepers, those who underestimate their sleep duration (similarly to our insomnia participants at T1) displayed higher EEG beta power and lower EEG delta power during NREM sleep than those who correctly estimate their sleep duration^28^. Cervena and collaborators (2004) reported increased delta activity and decreased sigma and beta activity after CBTi in 9 participants with psychophysiological insomnia^55^, but the relationship with sleep perception post-CBTi was not assessed. The neurophysiological underpinnings of this resolution of sleep-state misperception and whether a psychological intervention could impact the EEG delta and sigma ratio remain to be further investigated.

In addition to improving sleep satisfaction and reducing sleep-state misperception, the benefits of CBTi have also been found to extend to quality of life^3^ and daytime functioning, including large effects on concentration, productivity and sleepiness^32^. Here, CBTi did not significantly improve subjective assessment of cognitive function when using the SASCI questionnaire. While we found no change in the TX group compared to WL, there was a trend for an improvement of self-reported cognitive function when the data were pooled post-CBTi (*P* =.079, medium effect size *g’*=.375). This result differs from the large effect size found by Kyle and collaborators (2020)^19^ when using the BC-CCI questionnaire^59^. The absence of significant improvement in our study could be explained by the fact that the SASCI-Q^41^ assesses more specific cognitive impairment domains with precise examples rather than the overall cognitive dissatisfaction. Yet, we showed that the improvement in SASCI-Q was associated with the decrease in insomnia severity (i.e., ISI score) in the TX group only. Our findings converge with the results of Kyle and collaborators, who showed that the effect of digital CBTi on subjective cognitive function was mostly mediated by insomnia severity^19^.

As expected, we did not find an effect of CBTi on attentional and working memory abilities (i.e., using ANT and N-back tasks), which is similar to what has been previously found^19^. While daytime impairment, especially cognitive function impairment such as difficulty focusing, memorizing and cognitive fatigue in general, is a major complaint in chronic insomnia^32,60^, standardized objective tests often fail to detect such difficulties. Comprehensive neuropsychological test batteries of cognitive function failed to show consistent objective cognitive impairment when comparing participants with insomnia to good sleepers despite daytime function complaints^61–63^. Specifically, for tasks similar to the ones used in our study, differences in accuracy and reaction times are inconsistently found using the ANT^64,65^ or tasks assessing working memory performance such as the N-back^66–70^. A small effect size was found in previous studies when comparing executive functioning between individuals with chronic insomnia and healthy good sleepers^44,71,72^. Interestingly, the only domain that was found improved post-CBTi (pooled analyses) was executive function, however, the effect was present in both (TX and WL) groups with similar effect sizes (g’=0.52 and g’=0,43 respectively). This suggests an effect of practice (i.e., learning effect) rather than a direct effect of CBTi, which is consistent with the effect of practice that has been previously reported in both young and older participants in the ANT^73,74^. The absence of objective improvement after CBTi in both attentional and working memory performance may also be explained by the fact that simple computerized tasks such as the ANT and N-back may not be sensitive to overall cognitive changes, compared to more ecological tasks requiring the recruitment of multiple cognitive domains.

Despite its merits, such as the use of a randomized controlled design (i.e., immediate treatment versus wait-list), the possibility to pool data post-CBTi as well as several subjective and objective measures to investigate efficacy of CBTi on sleep and cognition, some limitations of this study affect the interpretability of the findings. First, our sample size was relatively limited, especially for some of the secondary variables (e.g., sleep diaries, N-back task – see Supplementary Methods). The lack of power as a result of variability in attrition and difference in sample size depending on the variables may have affected our ability to detect some significant differences between timepoints as some effects emerged from the larger pooled sample only (i.e., pooled data post-CBTi), especially for sleep diaries-determined measures. While we found no difference in sex between participants who did or did not provide measures at T2 or post-CBTi, we observed that younger participants (30-40 years old) were more likely to drop-out during the wait period. Inactive wait-list groups are considered to be an efficient control of time and placebo effect to an active intervention compared to ‘No Treatment’^75^, however, it might have impacted the motivation to remain in the study. To retain motivation, future studies might consider using an active control group who undertake an intervention that have been proved to yielded small-to-moderate effect size on insomnia severity (e.g., self-guided relaxation training, exercise)^76,77^. Moreover, the variability in attrition highlights the difficulties faced with such demanding and complicated trials in chronic insomnia. Finally, it is also important to note that the extractions of sleep parameters from a single night of PSG at each time point may also be considered as a limitation: as sleep parameters may vary across nights within a same individual and between different individuals^78^, this might explain the group differences observed at baseline in some sleep parameters, despite randomization.

In summary, using a randomized controlled design including both self-reported and objective measures of sleep and cognition, the present findings confirm the efficacy of CBTi on self-reported measure of sleep quality rather than on objective changes in sleep architecture. In addition, CBTi reduced sleep misperception in sleep onset latency and sleep duration. Finally, no effect of CBTi on cognitive function was observed in the present trial. Our results however suggest that such improvements might only be seen in those whose subjective sleep quality is improved, as changes in self-reported sleep and cognition were correlated. Further studies are needed to assess the effects of CBTi on sleep microarchitecture (e.g., spindles, slow waves), and their relationships with changes in sleep perception and cognitive performance.

## Data Availability

All data produced in the present study are available upon reasonable request to the authors

## ACKNOWLEDGMENTS

We acknowledge the contributions of the following students who assisted in participants’ recruitment, data collection and data preprocessing: Jennifer Suliteanu, Kazem Habibi, Brian Hodhod, Alex Hillcoat, Kajamathy Subramaniam, Elizaveta Frolova, Emma-Maria Phillips, Rachel Hu, Loren Bies, Meaghan Pawlowski, Aminata Baldé, Alexandros Hadjinicolaou, Elissa Pierre, Victoria Yue, Laurence Vo Buu, Maryam Aboutiman, Shira Azoulay and all the volunteers. We acknowledge the contribution of the psychologists who provided CBTi, the research coordinators, and nurses. We also thank our sleep technologists Madeline Dickson and Elinah Mozhentiy, and Liza Perez from the Clinique SomnoMed for their contribution to the setup of sleep recordings. Finally, we would like to thank the participants for giving their time and energy into this research study.

## LIST OF ABBREVIATIONS

AHI: Apnea-Hypopnea Index
ANOVA: Analysis Of Variance
ANT: Attention Network Task
CBTi: Cognitive Behavioral Therapy for insomnia
HLM: Hierarchical Linear Modeling
ISI: Insomnia Severity Index
MoCA: Montreal Cognitive Assessment
NR: Night Review
NREM: Non-Rapid Eye Movement
PSG: polysomnography
PSQI: Pittsburgh Sleep Quality Index
REM: Rapid Eye Movement
RT: Reaction Time
SASCI-Q: Sahlgrenska Academy Self-reported Cognitive Impairment Questionnaire
SD: Standard Deviation
SE: Sleep Efficiency
SEM: Standard Error of the Mean
SL: Sleep Latency
SOL: Sleep Onset Latency
SSM: Sleep-State Misperception
TiB: Time in Bed
TSP: Total Sleep Period
TST: Total Sleep Time
TX: Treatment group
WASO: Wake After Sleep Onset
WL: Wait-List group
1YFU: one-year follow-up

