## Supplemental for "Effects of cognitive behavioral therapy for insomnia on subjective and objective measures of sleep and cognition"

Perrault et al., SLEEP

### SUPPLEMENTARY METHODS

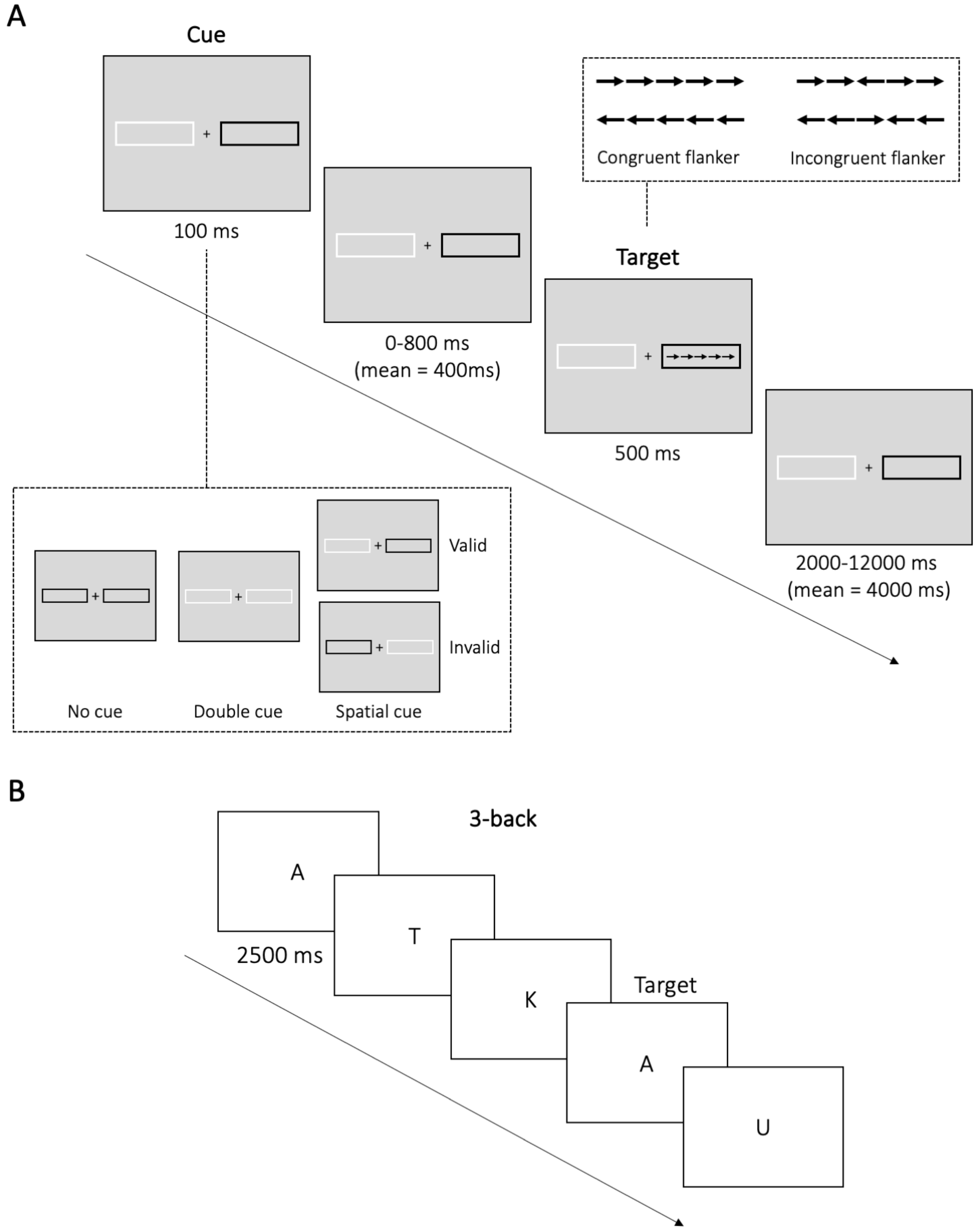


Supplementary figure 1 – Design of the cognitive tasks

*(A) In the **Attention Network Task (ANT)**, participants had to respond to the direction of the center arrow (target) of an array of 5 arrows (flanker arrows) for a total of 144 trials. The array of arrows appeared on either side of the screen and were cued by a flashing box. Cues were presented 0-800 ms (mean 400 ms) prior to the presentation of the arrows, highlighting the location of the array of arrows and were of three different types: no cue, double cue, or single cue which could validly be cueing the location of the array of arrows or not. The direction of the flanker arrows was either congruent or incongruent with the center (target) arrow.*

*(B) In the **N-back task**, a series of letters was presented at a fixed frequency (2500 ms), and participants were asked to press a button each time the current stimulus was identical to the stimulus presented N-letters before. We assessed three different difficulty levels: 1-, 2- and 3-back, each level comprising 45 trials (including 15 target trials) for a total of 135 trials. Accuracy and reaction times were extracted across all difficulty levels, as well as for each difficulty level separately.*

#### Treatment fidelity and participation attrition

##### Questionnaires

*Pittsburgh Sleep Quality Index (PSQI)* – At 3-months post-randomization (T2), of 33 participants in the TX group at T1, 29 (87.9%) filled the PSQI and 20 (68.9%) out of the 29 participants in the WL group filled the PSQI. There was no difference in gender (P > .05) or age (P > .05) between Groups at T1 (P > .05). However, there was an age difference between participants who did or did not provide T2 measures in the TX group, with participants who did not provide measures at T2 being older (63.5 years old) than the participants who did provide measures (46.8 years old; t= 3.29, P =.011). There was no difference in gender (P > .05) at T2 in the WL group. There was no significant difference in age or gender between participants who did or did not provide T2 measures in the WL group (P >.05).

For the pooled data, of the 62 participants who filled the PSQI Pre-CBTi, 54 (87.1%) filled the PSQI after completion of the CBTi program. There were no differences in gender or age between participants who did or did not provide measures at post-treatment (P > .05).

*Sahlgrenska Academy Self-reported Cognitive Impairment Questionnaire (SASCI-Q)* – At 3-months post-randomization, 27 (81.8%) out of 33 participants in the TX group filled the SASCI-Q at T2 and 20 (68.9%) out of the 29 participants in the WL group filled the SASCI-Q at T2. There was no difference in gender (P > .05) or age (P > .05) between Groups at T1 (P > .05). There was no difference in gender (P > .05) or age (P > .05) between groups at T2 (P > .05). Similarly, there was no age or gender difference between participants who did or did not provide T2 SASCI-Q measures in the TX or WL group (all P >.05).

For the pooled data, of the 62 participants who filled the SASCI-Q Pre-CBTi, 48 (77.4%) filled the SASCI-Q after completion of the CBTi program There were no differences in gender or age between participants who did or did not provide measures at post-treatment (P > .05).

##### Sleep diaries

At 3-months post-randomization, of 31 participants in the TX group at T1, 23 (74.1%) filled a sleep diary at T2 and 20 out of the 24 participants (83%) in the WL group filled a sleep diary at T2. There was no difference in gender (Fisher Exact Test) or age (student t-test) between Group at T1 (P > .05) and between participants who did or did not provide T2 measures in CBT and WL group (all P > .05).

For the pooled data, of the 55 participants who filled a sleep diary Pre-CBTi, 39 (70.9%) filled a sleep diary after completion of the CBTi program. There were no differences in gender or age (all P > .05) between participants who did or did not provide measures at post-treatment.

##### Polysomnographic recordings

At 3-months post-randomization, 27 (84.3%) out of the 32 participants in the TX group at T1 came back to the lab at T2 and 22 out of the 27 participants (81.4%) in the WL group came back to the lab at T2. There was no difference in gender (Fisher Exact Test) or age (student t-test) between Group at T1 (P > .05). We found no difference in age and gender between participants who did or did not provide T2 measures in the TX group (P > .05) but there was a significant difference in age in the WL group, with participants who did not provide measures at T2 being younger (36 ± 18 years old) than the participants who did provide measures (57 ± 10 years old; t= 17.7, P <.001).

For the pooled data, of the 59 participants who performed a sleep assessment Pre-CBTi, 46 (77.9%) came back to the lab after completion of the CBTi program. There were no differences in gender (P > .05) between participants who did or did not provide measures at post-treatment. There was a significant difference in age, with participants who did not provide measures Post-CBT being younger than the participants who did provide measures (Post-CBT: t= 23.9, P =.016).

Morning cognitive tasks

*Attention Network Task* - At 3-months post-randomization, of 32 participants in the TX group at T1, 26 (81.3%) completed the ANT at T2 and 21 (72.4%) out of the 29 participants in the WL group completed the ANT at T2. There was no difference in gender (P > .05) or age (P > .05) between Group at T1 (P > .05). There was no difference in gender (P > .05) or age (P > .05) between groups at T1 (P > .05). Similarly, there was no age or gender difference between participants who did or did not provide ANT measures at T2 in the TX or WL group (all P >.05).

For the pooled data, of the 61 participants who completed the ANT at baseline, 47 (77%) completed the ANT after completion of the CBTi program. There were no differences in gender or age between participants who did or did not provide measures at post-treatment (P > .05).

*N-back task* – At 3-months post-randomization, of 29 participants in the TX group at T1, 24 (82.8%) completed the N-back task at T2 and 18 (64.3%) out of the 28 participants in the WL group completed the N-back task at T2. There was no difference in gender (P > .05) or age (P > .05) between Group at T1 (P > .05). There was no difference in gender (P > .05) or age (P > .05) between groups at T1 (P > .05). Similarly, there was no age or gender difference between participants who did or did not provide N-back measures at T2 in the TX or WL group (all P >.05).

For the pooled data, of the 57 participants who completed the N-back task Pre-CBTi, 40 (70.2%) completed the N-back task after completion of the CBTi program. There were no differences in gender or age between participants who did or did not provide measures at post-treatment (P > .05).

### SUPPLEMENTARY RESULTS

**Table S1 - Insomnia severity trajectory (HLM)**

| | | INS (pooled) | | | |
|---|---|---|---|---|---|
| **Outcome measure** | | **Beta** | **SE** | **t** | **p** |
| ISI score (/28) | Pre | | | | |
| | Post | -7.8 | 0.63 | -12.3 | <.001 |
| | 1YFU | -8.2 | 0.65 | -12.6 | <.001 |

*ISI, insomnia severity index*

**Table S2 - Sleep measures (Pre versus Post-CBTi)**

| Outcome measure | (N) | | INS (pooled) Mean | (SD)¥ | Cohen g' | Time F | P | Q |
|---|---|---|---|---|---|---|---|---|
| ISI score (/28) | (56) | | | | | | | |
| | | Pre | 17.30 | (3.7) | 1.698 | **85.13** | **<0.001*** | - |
| | | Post | 9.50 | (5.1) | | | | |
| PSQI score (/21) | (54) | | | | | | | |
| | | Pre | 10.3 | (2.8) | -1.270 | **40.16** | **<0.001*** | - |
| | | Post | 6.6 | (3.1) | | | | |
| **Sleep diaries** | (37) | | | | | | | |
| Mean SOL (min) | | Pre | 34.3 | (3.8) | 0.548 | **5.92** | **0.017*** | **0.034*** |
| | | Post | 22.8* | (2.8) | | | | |
| Mean TiB (min) | | Pre | 495.8 | (10.5) | 0.465 | **4.77** | **0.032*** | **0.038*** |
| | | Post | 467.9* | (7.7) | | | | |
| Mean TST (min) | | Pre | 343.4 | (12.8) | -0.296 | 2.16 | 0.146 | - |
| | | Post | 366.2 | (11.4) | | | | |
| Mean WASO (min) | | Pre | 71.3 | (9.4) | 0.439 | **4.82** | **0.031*** | **0.038*** |
| | | Post | 45.0* | (9.8) | | | | |
| Mean SE (%) | | Pre | 70 | (2.4) | -0.624 | **9.85** | **0.002*** | **0.012*** |
| | | Post | 78.8* | (2.1) | | | | |
| Mean Sleep Quality (/5) | | Pre | 2.9 | (0.1) | -0.577 | **6.21** | **0.015*** | **0.034*** |
| | | Post | 3.3* | (0.1) | | | | |
| **Polysomnographic recording (PSG)** | (46) | | | | | | | |
| TiB (min) | | Pre | 476.8 | (58.5) | 0.379 | 3.46 | 0.066 | - |
| | | Post | 453.4 | (62.6) | | | | |
| TST (min) | | Pre | 364.4 | (69.9) | 0.138 | 0.56 | 0.457 | - |
| | | Post | 354.9 | (65.4) | | | | |
| WASO (min) | | Pre | 78.3 | (44.6) | 0.091 | 0.22 | 0.641 | - |
| | | Post | 73.1 | (65.8) | | | | |
| SOL (min) | | Pre | 21.3 | (29.1) | 0.136 | 0.45 | 0.503 | - |
| | | Post | 17.6 | (23.4) | | | | |
| SL to consolidated NREM (min) | | Pre | 39.9 | (52.3) | 0.305 | 3.22 | 0.076 | - |
| | | Post | 24.5 | (25.4) | | | | |
| SE (%) | | Pre | 76.4 | (11.3) | -0.202 | 1.12 | 0.292 | - |
| | | Post | 78.9 | (13.3) | | | | |
| W%tsp | | Pre | 17.9 | (10.7) | 0.130 | 0.46 | 0.499 | - |
| | | Post | 16.3 | (13.2) | | | | |
| N1%tsp | | Pre | 4.1 | (2.4) | 0.093 | 0.21 | 0.648 | - |
| | | Post | 3.9 | (2.4) | | | | |
| N2%tsp | | Pre | 38.2 | (8.7) | -0.110 | 0.31 | 0.578 | - |
| | | Post | 39.4 | (11.2) | | | | |
| N3%tsp | | Pre | 18.8 | (6.8) | -0.078 | 0.18 | 0.674 | - |
| | | Post | 19.5 | (8.8) | | | | |
| REM%tsp | | Pre | 21.0 | (6.1) | -0.004 | 0.01 | 0.981 | - |
| | | Post | 21.0 | (6.7) | | | | |
| **Subjective night review (in-lab)** | | | | | | | | |
| TST (min) | (44) | | | | | | | |
| | | Pre | 313.0 | (82.4) | -0.341 | 2.85 | 0.095 | - |
| | | Post | 341.9 | (84.4) | | | | |
| SOL (min) | | | | | | | | |
| | | Pre | 43.8 | (60.5) | 0.188 | 0.80 | 0.372 | - |
| | | Post | 32.8 | (54.3) | | | | |
| **SSM (PSG vs night review)** | | | | | | | | |
| SSM TST (min) | (44) | | | | | | | |
| | | Pre | -56.3 | (67.7) | -0.541 | **6.95** | **0.009*** | - |
| | | Post | -16,3* | (77) | | | | |
| SSM SOL (min) | | | | | | | | |
| | | Pre | 23.0 | (53.9) | 0.128 | 0.38 | 0.537 | - |
| | | Post | 15.8 | (56.3) | | | | |

*¥ SD: Standard Deviation or SEM for sleep diaries*

*ISI, insomnia severity score; PSQI, Pittsburgh sleep quality index; SOL, sleep onset latency; TiB, time in bed; TST, total sleep time; WASO, wake after sleep onset; SE, sleep efficiency; SL, sleep latency; NREM, non-rapid eye movement; TSP, total sleep period; REM, rapid eye movement; SSM, sleep-state misperception*

**Table S3 - Cognitive measures (Pre vs Post-CBTi)**

| Outcome measure | (N) | INS (pooled) Mean | (SD) | Cohen g' | Time F | *P* | *Q* |
|---|---|---|---|---|---|---|---|
| **<u>Subjective cognitive functioning</u>** | (43) | | | | | | |
| SASCI-Q score (/203) | Pre | 106.5 | (36.8) | 0.375 | 3.16 | 0.079 | - |
| | Post | 93.8 | (35.5) | | | | |
| **<u>Objective cognitive functioning</u>** | | | | | | | |
| <u>ANT</u> | (44) | | | | | | |
| Accuracy (% correct) | Pre | 87 | (20) | -0.532 | **5.74** | **0.017*** | **0.042*** |
| | Post | 92.3* | (11) | | | | |
| Reaction time - correct (ms) | Pre | 665 | (83.1) | 0.178 | 0.70 | 0.406 | |
| | Post | 653 | (87.1) | | | | |
| Alerting (ms) | Pre | 26 | (28) | -0.297 | 1.94 | 0.168 | |
| | Post | 35 | (30) | | | | |
| Orienting (ms) | Pre | 88 | (38) | -0.094 | 0.19 | 0.661 | |
| | Post | 92 | (37) | | | | |
| Executive function (ms) | Pre | 112 | (28) | 0.540 | **6.41** | **0.013*** | **0.042*** |
| | Post | 97.1* | (26) | | | | |
| <u>N-back</u> | (40) | | | | | | |
| Accuracy (% correct) | Pre | 88.4 | (6.4) | -0.379 | 2.87 | 0.094 | - |
| | Post | 90.3 | (5.5) | | | | |
| Accuracy - 1-back (%) | Pre | 95.9 | (6.7) | -0.291 | 1.70 | 0.197 | - |
| | Post | 97.3 | (3.3) | | | | |
| Accuracy - 2-back (%) | Pre | 87.8 | (9.4) | -0.410 | 3.35 | 0.071 | - |
| | Post | 90.9 | (7.6) | | | | |
| Accuracy - 3-back (%) | Pre | 81.4 | (7.5) | -0.207 | 0.86 | 0.358 | - |
| | Post | 82.9 | (8.1) | | | | |
| Reaction time (ms) | Pre | 723 | (156) | 0.157 | 0.49 | 0.485 | - |
| | Post | 703 | (136) | | | | |
| RT - 1-back (ms) | Pre | 601 | (136) | 0.052 | 0.05 | 0.817 | - |
| | Post | 595 | (156) | | | | |
| RT - 2-back (ms) | Pre | 783 | (192) | 0.199 | 0.80 | 0.375 | - |
| | Post | 749 | (180) | | | | |
| RT - 3-back (ms) | Pre | 828 | (217) | 0.233 | 1.08 | 0.301 | - |
| | Post | 776 | (229) | | | | |

*SASCI-Q, Sahlgrenska academy self-reported cognitive impairment questionnaire; ANT, attention network task*
*RT, reaction time*
